# PONT: A Protocol for Online Neuropsychological Testing

**DOI:** 10.1101/2021.05.27.21257953

**Authors:** William Saban, Richard B. Ivry

## Abstract

A major challenge for neuropsychological research arises from the fact that we are dealing with a limited resource: The patients. Not only is it difficult to identify and recruit these individuals, but their ability to participate in research projects can be limited by their medical condition. As such, sample sizes are small and considerable time (e.g., 2 yrs) is required to complete a study. To address limitations inherent to lab-based neuropsychological research, we developed a protocol for online neuropsychological testing (PONT). We describe the implementation of PONT and provide the required information and materials for recruiting participants, conducting remote neurological evaluations, and testing patients in an automated, self-administered manner. The protocol can be easily tailored to target a broad range of patient groups, especially those that can be contacted via support groups or multi-site collaborations. To highlight the operation of PONT and describe some of the unique challenges that arise in on-line neuropsychological research, we summarize our experience using PONT in a research program involving individuals with Parkinson’s disease and spinocerebellar ataxia. In a 10-month period, by contacting 646 support group coordinators, we were able to assemble a participant pool with over 100 patients in each group from across the United States. Moreover, we completed six experiments (n>300) exploring their performance on a range of tasks examining motor and cognitive abilities. The efficiency of PONT in terms of data collection, combined with the convenience it offers the participants, promises a new approach that can increase the impact of neuropsychological research.

## Introduction

Understanding the functional organization of the brain requires the use of converging methods of exploration. The spatial resolution of functional magnetic resonance imaging (fMRI) and temporal resolution of electroencephalography (EEG) can reveal the dynamics of computations in local regions, as well as coordination of activity across neural networks. A limitation with these methods is that they are fundamentally correlational, revealing the relationship between cognitive events and brain regions. Traditionally, perturbation methods such as optogenetics or lesion studies, have been seen as approaches that provide stronger tests of causality. Lesion studies, such as neuropsychological studies on patients with focal brain pathology, could provide us with causal evidence for the potential role of a specific brain region in a cognitive function.

Empirical investigations on patients with brain pathology have been central in the emergence of behavioral neurology as an area of specialization, and, with its more theoretical focus, cognitive neuroscience. Information obtained through neuropsychological testing has been fundamental in advancing our understanding of the nature of brain-behavior relations (Luria, 1966) with important clinical implications (Lezak, 2000). Neuropsychological testing is not only important in evaluating the cognitive changes associated with focal and degenerative disorders of specific brain regions but has also been an essential tool in determining the contribution of targeted brain regions and particular cognitive operations. For example, neuropsychological observations from the 19^th^ century led to the classic “Broca-Wernicke-Lichtheim-Geschwind” model of language function (Geschwind, 1970; Poeppel, 2014).

### Challenges with neuropsychological testing

While neuropsychological research has been a cornerstone for cognitive neuroscience, its relative importance to the field has diminished over the past generation with the emergence of new technologies. This point is made salient when scanning the table of contents of the journals of the field: Papers using fMRI and EEG dominate the publications, with only an occasional issue including a study involving work with neurological patients. For example, in PubMed searches restricted to papers since 2011, the key word “fMRI” yielded 409,310 hits, whereas the key word “neuropsychological” yielded only 72,835 hits.

There are several methodological issues that constrain the availability and utility of neuropsychological research. First, the studies require access to patients who have relatively homogenous neurological pathology. This is quite challenging when drawing on patients with focal insults from stroke or tumor, or when studying rare degenerative disorders. As such, the sample size in neuropsychological research tends to be small, such as 8 participants (Casini & Ivry, 1999). Often, researchers have very limited access to these hard-to-reach populations, especially with rare disorders such as spinocerebellar ataxia (SCA). Second, given that the recruitment is usually from a clinic or local community, the sample may not be representative of the population; for example, samples may be skewed if based on patients who are active in support groups. Third, given their neurological condition, the patients may have limited time and energy. Their disabilities make it challenging to recruit the participants to come to the lab, and for those that do, the experiments have to be tailored to avoid taxing the participants’ mental capacity. As a result, it can take quite a bit of time to complete a single experiment, let alone a package of studies that might make for a comprehensive story. Hence, the progress and efficiency of each study can be very restricted.

### Addressing the challenges with neuropsychological testing

One solution that we have pursued in our research on the cerebellum has been to take the lab into the field, setting up a testing room at the annual meeting of the National Ataxia Foundation. This conference is patient-focused, providing people with ataxia a snapshot of the latest findings in basic research and clinical interventions, and an opportunity to share their experiences with their peer group. For the past decade, we have set up a testing room at the conference and, over a 3-day period, been able to test about 15 people, a much more efficient way to complete a single study compared to the more traditional approach of enlisting ataxia patients in the Berkeley community to come to the lab. However, this approach is not ideal for multi-experiment projects, and entails considerable cost to send a team of researchers required to recruit the participants and coordinate the testing schedule. Conferences in which researchers are reaching out to patients are still a slow and expensive solution.

An alternative and simpler solution might come from the use of the internet. In recent years, behavioral researchers in different domains of studies have developed online protocol s to reach larger (Adjerid & Kelley, 2018) and more diverse (Casler, Bickel, & Hackett, 2013) populations than feasible with lab-based methods. Within psychology, platforms such as Amazon Mechanical Turk have been used to efficiently collect behavioral data (Crump, 2013). A number of studies have shown that the data obtained in online testing is as reliable and valid as in-person testing (Buhrmester, Kwang, & Gosling, 2011; Casler, Bickel, & Hackett, 2013; Chandler & Shapiro, 2016). Clinical psychology (Chandler & Shapiro, 2016) and developmental studies (Tran, Cabral, Patel, & Cusack, 2017), which have more limited in-person access to their research population than other domains, have particularly benefited because of the unique challenges that can arise in recruiting these populations to the lab.

To date, online protocols have been used in a limited manner in medical (Ranard et al., 2014) and neuropsychological research, with the focus on collecting survey data (e.g., Gong et al., 2020) or patient recruitment (Hurvitz, Gross, Gannotti, Bailes, & Horn, 2020). Here we report an online protocol designed to recruit participants and administer behavioral tests for neuropsychological research. While we developed this protocol to facilitate our research program involving patients with subcortical degenerative disorders, the protocol can be readily adopted for different populations. As such, it provides a valuable and efficient new approach to conduct neuropsychological research and, we hope, will contribute to the neurology, neuropsychology, and cognitive neuroscience communities.

## PONT

We call the new protocol PONT, an acronym for Protocol for Online Neuropsychological Testing. The PONT protocol entails a comprehensive package that addresses challenges involved in recruitment, neuropsychological evaluation, behavioral testing, and administration to support neuropsychological research. In the initial ten-month period, we contacted 646 support group coordinators to recruit large samples of individuals with degenerative disorders of the cerebellum (SCA) or basal ganglia (Parkinson’s disease (PD)). We established a workflow for online recruitment, neuropsychological assessment, behavioral testing, and follow-up. During this ten-month period, we have completed six experiments testing motor and cognitive abilities, and report one of these studies in this paper.

## Methods

### How does PONT work?

PONT entails five primary steps (Figure 1): 1) Contacting support group leaders to advertise the project; 2) Having interested individuals initiate contact with us, a requirement set by our IRB protocol; 3) Conducting interactive, remote neuropsychological assessments; 4) Automated administration of the experimental tasks; 5) Obtaining feedback and providing payment. A detailed, step-by-step, description of the protocol is available at this <u>online PONT-general workflow link</u> (full URL address is provided in the Appendix). The protocol was approved by the institutional review board at the University of California, Berkeley.

**Figure 1.**
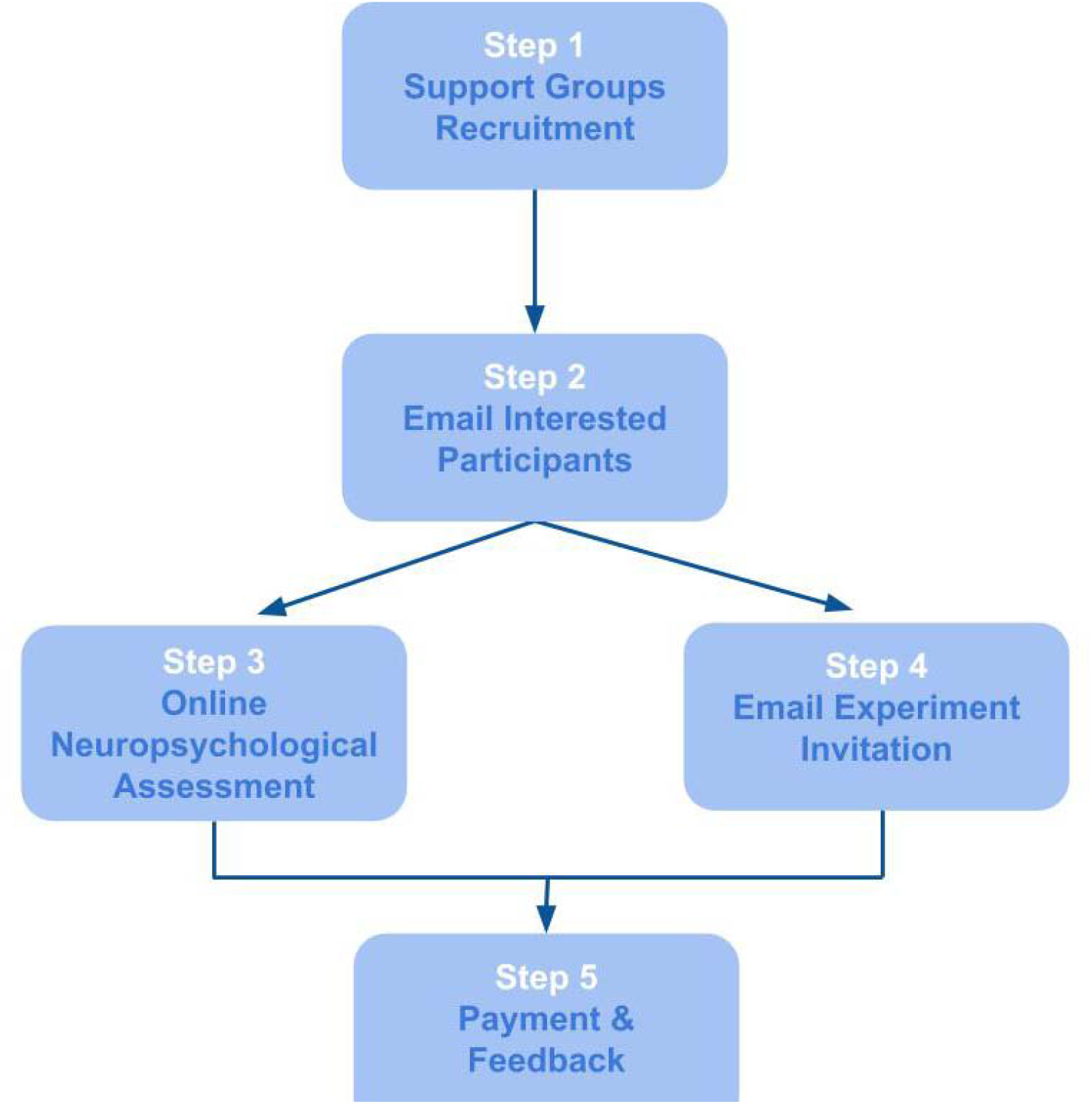
General PONT workflow.

#### 1. Working through support group leaders

Our IRB recruitment rules do not allow UC Berkeley researchers to directly contact potential participants; rather, we can provide descriptive materials with information indicating how interested individuals can contact us. Given this constraint, we sent emails to 646 contact address of support groups across the USA. This information was available on the National Ataxia Foundation website and the Parkinson and Movement Disorder Alliance website. The initial email provides a description of PONT and provides text that the support group administrators can pass along to their group members. Although we don’t explicitly request a response from the administrators, our experience is that those who pass along our recruitment information inform us of this in a return email. The response rate is modest: Following our initial email and a follow-up two weeks later to those who haven’t responded, we estimate that approximately 15% of the support groups forward our information to their members. While this number is low, it does mean that our recruitment information has reached the membership of around 100 support groups.

This procedure is a variant of snowball sampling (Goodman, 1961), a method that has been employed to recruit patients and other hard-to-reach groups for clinical studies. Here we modify this procedure to enlist participants via support groups and, cascading from there, word of mouth for our behavioral studies. The materials used in this phase of the recruitment process can be found at this <u>online Outreach Forms link</u> (Step #1; full URL address is provided in the Appendix). These can be readily adopted by any research group that is targeting a population associated with support organizations and web-based groups, or when the sample might be enlisted as part of a multi-lab collaborations.

#### 2. Enrollment of the participants

In the second step, individuals who received our flyer via their support group leaders and wished to participate could register for the study either by emailing the lab or filling out this <u>online Registration Form link</u> (full URL address is provided in the Appendix). In this way, the participant-initiated contact with the research team, a requirement set forth in our IRB contact guidelines. Between November 2019 and October 2020, 103 individuals with SCA (age range: 24 to 88) and 133 individuals with PD (age range: 48 to 83) enrolled in the project. Reflecting the distribution of the support groups, this sample included individuals that are currently residing in 30 states, such as Hawaii, Kentucky, Ohio, California and more. Thus, our sample for PONT is much more geographically diverse than would occur in a typical laboratory-based study.

Control participants were recruited via advertisements posted on the Craigslist website. The advertisement instructed interested individuals to complete our online form. The advertisement indicated that participation was restricted to individuals between the ages of 35 and 80. Over a 9-month period, we have enrolled 159 individuals in the control group.

#### 3. Online Interview: Demographics, Neuropsychological Assessment, and Medical Evaluation

The third step in PONT involved an online video interview that included a neurological and neuropsychological evaluation. The materials used in our interview and evaluation are provided at this <u>online Participant Interview link</u> (full URL address is provided in the Appendix). While the relevant instruments will vary depending on the interests of a research group, these documents demonstrate some of the modifications we adopted over the course of pilot work with PONT (see below).

Registered individuals were contacted by email to invite them to participate in an online, live interview with an experimenter. This session provided an opportunity to describe the overall objective of the project, confirm basic demographic information, perform a short assessment of cognitive status, and for the individuals with PD or SCA, obtain a medical history and abbreviated neurological exam. The invitation email indicated that the session would require that the participant enlist another individual (e.g., caregiver, family member) to join in for part of the session to assist with test administration and video recording. It also indicated that participation in the project would require the ability to use a computer and respond unassisted on a computer keyboard.

Approximately 240 individuals were invited for the on-line interview. If no response was received, a follow-up email was sent approximately 14 days later. We have received 125 affirmative responses over the past nine months. To date, 118 interviews have been conducted (60 PD, 18 SCA, 40 Controls).

At the start of the on-line session, the experimenter provided an overview of the mission of our PONT project and the goals of the interview. This introduction provided an opportunity to emphasize orally that participation is voluntary and involves a research project that will not impact the participant’s clinical care nor provide any direct clinical benefit. After providing informed consent, the participant completed a demographic questionnaire. The experimenter then administered the Montreal Cognitive Assessment test (MoCA, Nasreddine et al., 2005) as a brief evaluation of cognitive status. For control participants, the session ended with the completion of the MoCA.

The PD and SCA participants continued on to the medical evaluation phase. First, the experimenter obtained the participant’s medical history, asking questions about age at diagnosis, medication and other relevant information (e.g., DBS for PD; genetic subtype if known for SCA), and a screening for other neurological or psychiatric conditions. Second, the experimenter administered a modified version of the motor section of the Unified Parkinson’s Disease Rating Scale, (UPDRS; Goetz, 2003) to the PD participants and the Scale for Assessment and Rating of Ataxia (SARA, (Schmitz-Hübsch et al., 2006) to the SCA participants.

Modifications were made to these assessment instruments to make them more appropriate for online testing. For the MoCA test, we eliminated “Alternating Trail Making” since this would require that we provide a paper copy of the task. For the UPDRS and SARA, we modified items that require the presence of a trained individual to ensure safe administration. We eliminated the “Postural Stability task” from the UPDRS since it requires that the experimenter apply an abrupt pull on the shoulders of the participant. We modified three items on the UPDRS (“Arising from Chair”, “Posture”, and “Gait”), obtaining self-reports from the participant rather than the standard evaluation by the experimenter. Similarly, we obtained self-reports of stance and gait for the SARA rather than observe the participant on these items. For the self-reports, we provided the scale options to the participant (e.g., on the SARA item for gait, 0, normal/no difficulty, to 8, unable to walk even supported). The scores for the MoCA and UPDRS batteries were adjusted to reflect these modifications. For the online MoCA, the observed score was divided by 29 (the maximum online score), and then multiplied by 30 (the maximum score on the standard test). Hence, if a participant obtained a score of 26, the adjusted score will be (26/29)*30, or 26.9. The same adjustment procedure was performed for the UPDRS. No adjustment was required for the SARA.

The interview took around 30 minutes for the control participants and 40-60 minutes for the PD and SCA participants.

#### 4. Completion of Online Experiments

The experiments were programmed in Gorilla Experiment Builder (Anwyl-Irvine, Massonnié, Flitton, Kirkham & Evershed, 2018). For a given experiment, the participant was emailed an individual link that assigned a unique participant ID, providing a means to ensure that the data were stored in anonymized and confidential manner (see template at this <u>online Experiment Invite link</u>, Step #4; full URL address is provided in the Appendix). This email also included a brief overview of the experimental task (e.g., a study about how PD impacts finger movements or a study looking at mathematical skills in SCA). The email clearly stated that the participant should click on the link when they are ready to complete the experimental task, allowing sufficient time to complete the study (30 to 60 minutes depending on the experiment). In this manner, administration of the experimental task was entirely automated, with the participant having complete flexibility in terms of scheduling.

To date, we have run six experiments with the PONT protocol, with the studies looking at various questions related to the involvement of the basal ganglia and/or cerebellum in motor learning, language, and mathematics. For the experiments involving both patient groups, we set a recruitment goal of 20 individuals/group (including controls), although the exact numbers vary given that recruitment is done in batches of approximately 70 emails/group. The response rate to a given batch is approximately 15% and follow-up emails are sent after about two weeks to repeat the invitation. As such, it takes approximately four to eight weeks to complete a single experiment.

#### 5. Participant Feedback and Payment

After completing the online experiment, the participant is sent an automated thank you note and a short online form for providing feedback about the task (e.g., rate difficulty, level of engagement, see Table 2). In addition, we sent the participant an email with information about payment (see template at this <u>online Feedback Form</u>, Step #5). Reimbursement is at $20/hour and the participant can opt to be paid by check sent via regular mail, Paypal, or with an Amazon gift card. We also reimburse for the online, live interview session to further reinforce that this is a research project unrelated to their medical care. The experimenter monitors the feedback reports and, when appropriate, sends an email response to the participant.

The participant’s data is downloaded from the Gorilla protocol to a secure laboratory computer. Within this secure, local environment, we have the ability to link the unique participant ID code used to access the experiment with personal information. This allows us to build a database to track the involvement of each individual in PONT, as well as make comparisons across experiments.

### PONT in action: Sequence learning in PD and SCA

To demonstrate the feasibility and efficiency of PONT, we report the results from an experiment on motor sequence learning in which we used the discrete sequence production (DSP) task. On each trial, the participant produces a sequence of four keypresses in response to a visual display. We chose to use a motor sequence learning task for two reasons. First, there exists a large literature on the involvement of the BG and the cerebellum in sequence learning (Debas et al., 2010; Doyon et al., 2002; Jouen, 2013), including studies specifically on PD and SCA (Gamble et al., 2014; Roy, Saint-Cyr, Taylor, & Lang, 1993; Ruitenberg et al., 2016; Shin & Ivry, 2003a; R. M C Spencer & Ivry, 2009; Tremblay et al., 2010). Thus, we can compare the results from our online study with published work from traditional, in-person studies. Second, the requirement that a trial consist of four, sequential responses makes this a relatively hard task in terms of response demands compared to tasks requiring a single response (e.g., 2-choice RT). As such, we can evaluate the performance of our code across the range of hardware protocol s used by the participants, as well as evaluate participants’ performance on a relatively demanding motor task.

Skill acquisition has been associated with two prominent processes: 1) Memory retrieval that becomes enhanced with practice (Logan, 1988) and 2) improved efficiency in the execution of algorithmic operations (Tenison & Anderson, 2016). We modified the DSP task to look at how degeneration of the BG or cerebellum impact these two learning processes. We compared practice benefits for repeated items (memory-based learning) with practice benefits for non-repeated items (algorithm-based learning). While we expected the PD and SCA groups would be slower than the controls overall, we expected sequence execution time would become faster over the experimental session for all three groups. Our primary focus was to make group comparisons of the learning benefits for the repetition and non-repetition conditions, providing assays of memory-based and algorithm-based learning, respectively.

#### Participants

Drawing on the PONT participant pool, email invitations were initially sent to 85, 66, and 89 individuals in the Control, SCA, and PD groups, respectively, with the differences reflecting the pool size for each group at the time of the email. The overall response rate to this first email was around 20%. Follow-up emails were sent every few weeks, and after a few rounds, we reached our goal of a minimum of 20 participants per group. Of those who initiated the study, we excluded the data of three participants (one per group) who failed to respond correctly to the attention probes (see below), and four participants (1 Control, 2 SCA, 1 PD) who failed to complete the experiment (either aborting the program or a loss of internet connectivity during the session). The final sample of participants included in the analyses reported below was composed of 62 participants, 22 Control, 17 SCA, and 23 PD.

Table 1 provides demographic information for the three groups, as well as the adjusted MoCA, SARA (SCA), and UPDRS (PD) scores. The SCA group was composed of 12 individuals with a known genetic subtype and 5 individuals with an unknown etiology (idiopathic ataxia). The mean duration since diagnosis for the SCA group was 5.9 years (SE=1.8) and the mean SARA score was 9.1 (SE=.9, range: 2 - 14.5). The mean duration since diagnosis for the PD group was 6.5 years (SE=.9) and mean score on the motor section of the UPDRS was 16.2 (SE=.9, range: 9.7-24.8). None of the individuals in the PD group had undergone surgical intervention as part of their treatment (e.g., DBS) and all were tested while on their current medication regimen.

**Table 1.** Demographic and neuropsychological summary of all groups.

|  | Years of Education | # of Females | Age | MoCA | Motor assessment |
| --- | --- | --- | --- | --- | --- |
| **Control** | 17.7, [.4], (14-20) | 12 | 63.6, [2.3], (40-78) | 27.8, [.4], (21-30) |  |
| **SCA** | 17.1, [.8], (12-22) | 12 | 56.3, [2.6], (34-80) | 25.8, [.6], (22-29) | 9.1, [.9], (2-14.5) |
| **PD** | 17.2, [.5], (12-22) | 12 | 65.3, [1.5], (50-79) | 26.8, [.5], (23-30) | 16.2, [.9], (9.7-24.8) |

Mean, [SE], and (Range) for each demographic and neuropsychological variable. The scores of the MoCA and UPDRS are adjusted for online administration.

#### Procedure

The participant used their home computer and keyboard to perform the experiment. Given this and the fact that we did not control for viewing distance, the size of the stimuli varied across individuals. All of the instructions were provided on the monitor in an automated manner, with the program advancing under the participant’s control.

The participant was instructed to place his or her fingers (thumbs excluded) on the keyboard, using the keys “z”, “x”, “c”, “v” for the left hand and the keys “m”, “<“, “>“, and “?” for the right hand. Eight placeholders were displayed on the screen, with each placeholder corresponding to one of the keys of the keyboard in a spatially compatible manner (Figure 2).

**Figure 2.**
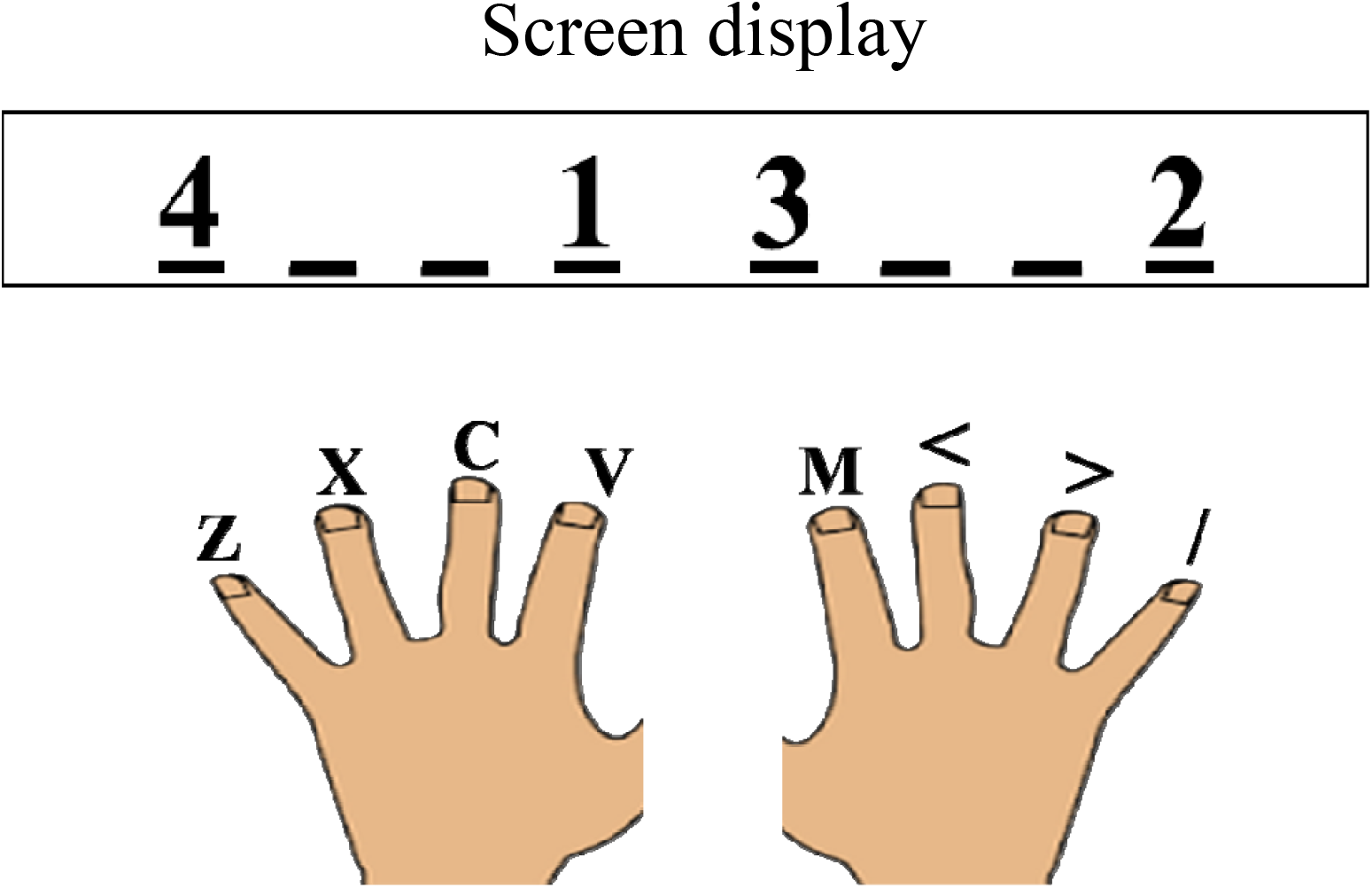
Each line on the screen corresponded to a finger position in a spatially compatible manner. The numbers indicate the order of the keypresses for the current trial. In this example, the participant should press left index (“v”), right pinky (“/”), right index (“m”), and left pinky (“z”).

At the start of each trial, a black fixation cross appeared in the middle of a white background. After 500 ms, the fixation cross was replaced by a stimulus display that consisted of the numbers, “1”, “2”, “3” and “4”, with each number positioned over one of the placeholders. The numbers specified the required sequence for that trial. In the example shown in Figure 2, the correct sequence required sequential key presses with the left index finger, right pinky, right index finger, and left pinky. The instructions emphasized that the sequence of responses should be performed as quickly and accurately as possible. If a keypress was not detected within 3000 ms for each element of a given sequence, the phrase “Respond faster” appeared for 200 ms. If a single response was not detected within 4000 ms or the wrong key was pressed, the trial was aborted. In the case of an erroneous keypress, a red “X” was presented above the placeholders. When the entire sequence was executed successfully, a green “√” appeared above the placeholders. The feedback screen remained visible for 500 ms, after which a fixation cross appeared for 500 ms indicating the start of the inter-trial interval.

To test memory-based and algorithm-based learning, we created two nonoverlapping categories of sequences. For memory-based learning, a set of repeating sequences was created, composed of eight 4-element sequences. For algorithm-based learning, a set of novel sequences was created, composed of 192, 4-element unique sequences. The sequences for each condition were determined randomly for each participant with the constraints that each sequence required at least one key press from each hand. In addition, in each sequence, a finger should not press a key more than once.

The experimental block was composed of 384 trials, divided into 24 blocks of 16 trials each. Each block included one presentation of each of the eight sequences in the repetition condition and eight of the unique sequences, with the order of the 16 sequences determined randomly within each block. A minimum one-minute break was provided after each run of four blocks, with the participant pressing the “v” key when ready to continue the experiment.

To ensure that participants remained attentive, we included five “attention probes” on the instruction pages that appeared in the start of the experiment or during the experimental block. For example, an attention probe might instruct the participant to press a specific key rather than selecting the “next” button on the screen to advance the experiment (e.g., “Do not press the “next” button. Press the letter ‘A’ to continue”). If the participant failed to respond as instructed on these probes, the experiment continued, but the participant’s results were not included in the analysis.

The experiment took approximately 45 minutes to complete.

## Results

Accuracy rates were 80%, 78%, and 78% for the Control, SCA, and PD groups, respectively (F(2,59)=.198, *p*=.821; one-way ANOVA). Most of the errors involved an erroneous keypress (16% of all trials), and the remaining errors were due to a failure to make one of the keypresses within 4 s (5% of all trials). We excluded trials in which participants failed to complete the full sequence within 4 s (3% of the remaining trials).

Our primary dependent variable of interest was execution time (ET), the time from the first key press to the time of the last key press. Before turning to these data, we considered potential trade-offs in performance. First, to determine if there was a speed-accuracy trade-off, we looked at the correlation between ET and accuracy for each group. These correlations were all negative (Control=-.14, SCA=-.69, PD=-.20), indicating that participants who made the most errors also tended to be the slowest in completing the sequence, the opposite pattern of a speed-accuracy trade-off. Second, we were concerned that participants might vary in the degree to which they prepared the series of responses prior to initiating the first movement. For example, a participant might opt to preplan the full sequence prior to making the first keypress or plan the responses in some sort of sequential manner. To look at possible trade-offs between pre-planning and execution time, we computed the correlation between RT (time to first keypress) and ET (which starts at the time of the first keypress). These correlations were all positive (Control=.75, SCA=.57, PD=.51), indicating no ET-preplanning tradeoff.

Figure 3 shows ET as a function of cycle of learning and group, with separate functions for the no-repetition and repetition conditions. Across all conditions, the SCA group was 362.8 ms slower than the Control group in executing the sequences (SE=164.2, *p*=.031). The PD group was 167.6 ms slower than the Control group, but this effect was not significant (SE=151.6, *p*=.274). To statistically evaluate learning, we employed a linear mixed-effect model with the factors Group, Repetition Condition, and Cycle, and participant as a random factor (R software, lme4 library, Bates, Mächler, Bolker, & Walker, 2015). For the Cycle variable, we averaged across pairs of blocks to collapse the 24 blocks into 12 cycles. We also included years of education, age, and MoCA score as covariates in the model.

**Figure 3.**
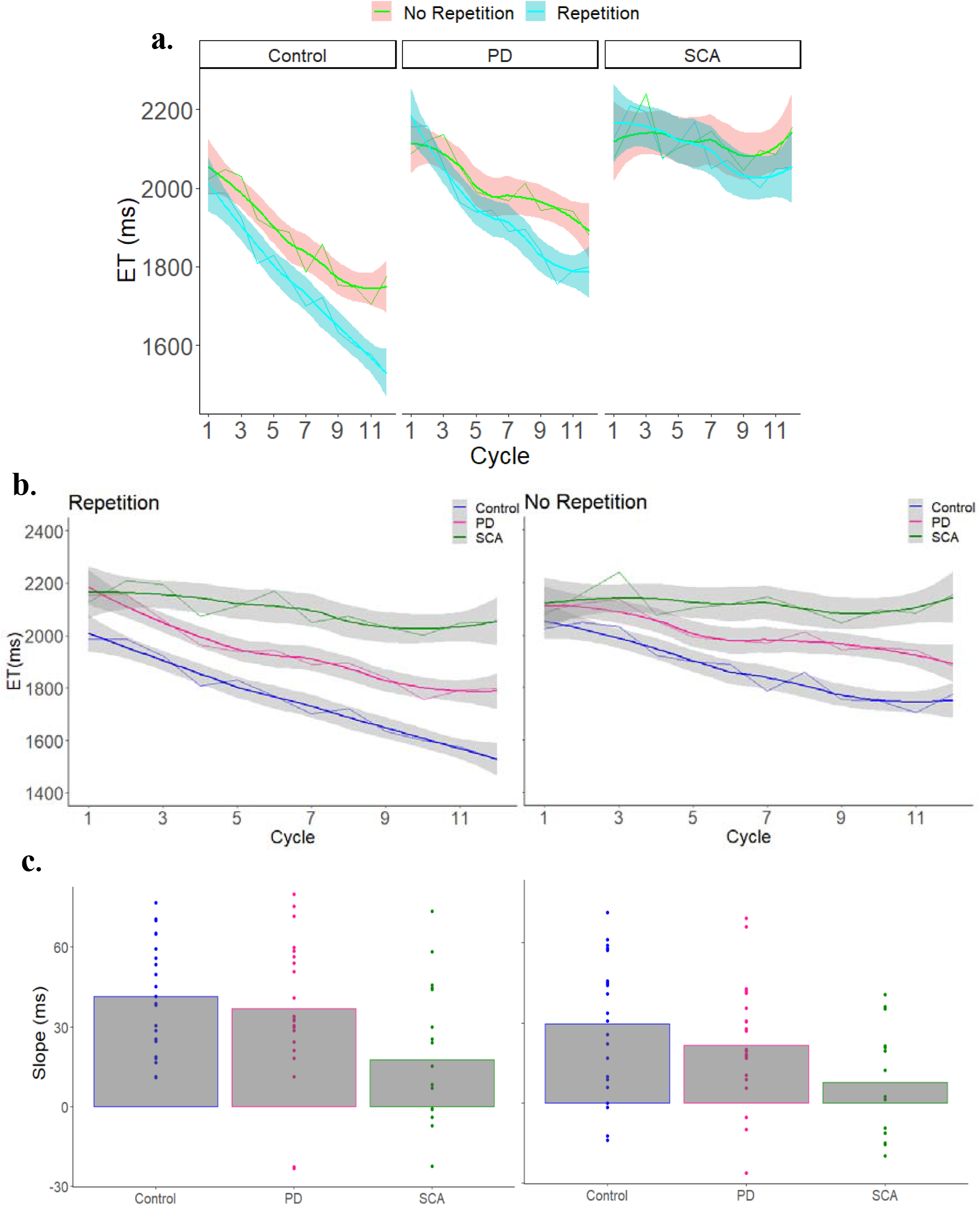
(**a**) ET as a function of cycle of learning, repetition condition, and group. (**b**) The effect of cycle of learning in each group for each repetition condition separately. (**c**) The slope for each individual (dots) in each repetition condition. Error bars = 95% confidence level.

The control participants got faster over the experimental session with an overall slope of -35.8 ms/cycle (SE=1.9, *p*<.0001). These participants showed significant learning in both the no-repetition condition (−29.4 ms/cycle, SE=2.8, *p*<.0001) and the repetition condition (−41.5 ms/cycle, SE=2.5, *p*<.0001). Similar to what has been observed with the DSP task in young adults (Jouen, 2013). the improvement was more pronounced for the repetition condition than for the no-repetition condition (difference in slope: -11.8 ms/cycle, SE=3.7, *p*=.001).

The patient groups also got faster over the experimental session with an overall slope of -29.2 ms/cycle for the PD group (SE=1.9, *p*<.0001) and -13.1 ms/cycle for the SCA group (SE=2.3, *p*<.0001). The improvement was significant in both the no-repetition condition (PD: - 20.5 ms/cycle, SE=2.7, *p*<.0001; SCA: -7.8 ms/cycle, SE=3.3, *p*=.019) and repetition condition (PD: -37.1 ms/cycle, SE=2.6, *p*<.0001; SCA: -18.4 ms/cycle, SE=3.0, *p*<.0001). Similar to the control participants, this magnitude of the improvement was more pronounced for the repetition condition than for the no-repetition condition (difference in slope: PD=-16.8 ms/cycle, SE=3.7, *p*=.001; SCA=-10.3 ms/cycle, SE=4.5, *p*=.023).

We next compared the learning effects for each patient group to the Control group. Both patient groups showed less improvement than the controls in the no-repetition condition, (SCA vs. Control: difference in slope: 21.6 ms/cycle, SE=4.19, *p*<.0001; PD vs. Control: difference in slope: 9.1 ms/cycle, SE=3.8, *p*=.017). However, only the SCA group was impaired in the repetition condition, showing less improvement than the Control group (23.2 ms/cycle, SE=4.1, *p*<.0001); the comparison between the Control and PD groups was not significant (4.1 ms/cycle, SE=3.7, *p*=.275). In a comparison of the two patient groups, the SCA group showed less improvement than the PD group on the no-repetition condition (12.6 ms/cycle, SE=4.23, *p*=.002) and repetition condition (18.7 ms/cycle, SE=3.94, *p*<.001). In terms of the covariates, there were no significant effects of education (18.5 ms/year, SE=67.9, *p*=.786), age (114.2/year, SE=71.1, *p*=.113), or MoCA score (−122.6/point, SE=74.2, *p*=.104).

Taken together, the results demonstrate the viability of using an automated, online protocol to examine sequence learning in neurological populations. As expected, the two patient groups were slower than the Control group overall, although this effect was only significant for the SCA group. More importantly, the results show sequence learning impairments in both groups, similar to what has been observed in laboratory studies (Gamble et al., 2014; Molinari et al., 1997; Roy et al., 1993; Ruitenberg, Duthoo, Santens, Notebaert, & Abrahamse, 2015; Ruitenberg et al., 2016; Shin & Ivry, 2003b; R. M C Spencer & Ivry, 2009; Tremblay et al., 2010). Interestingly, the SCA group was impaired in both the repetition and the no repetition conditions, a pattern suggestive of impairment in both memory-based and algorithm-based learning. In contrast, the PD group was only significantly impaired in the no repetition condition, a pattern suggestive of impairment in algorithm-based learning.

Sequence production and learning has been the focus of many previous studies involving both of these patient groups. We are unaware of any studies with either patient group using the DSP task with repetition versus no repetition manipulation, precluding direct, cross-experiment comparisons. Most of the prior work has been done with the serial reaction time task (SRTT; (Muslimović, Post, Speelman, & Schmand, 2007; Shin & Ivry, 2003a; Tremblay et al., 2010)), a method in which sequence learning is assessed by comparing blocks of trials in which a sequence of length n repeats in a cyclic manner (e.g., n=8 and a block involves 10 cycles) to blocks in which the stimuli are selected at random. As such, sequence learning is operationalized as reductions in reaction time for a repeating sequence relative to improvements on the random blocks.

Using this measure, individuals with cerebellar pathology, either from degeneration or focal lesions, consistently exhibit a pronounced learning deficit (Molinari et al., 1997; Tzvi et al., 2017), one that within the present framework, would be attributed to memory-based learning. One exception is a study (Spencer & Ivry, 2009) that observed normal learning when the responses were made directly to the stimulus positions. It may be that these “direct” cues either support the formation of distinct memory associations not impacted by cerebellar pathology (e.g., direct stimulus-response mappings, not requiring an intermediary mapping operation), or that the direct cues provide a boost to memory formation. One could consider the random blocks as a long “non-repeating” sequence. For example, it was demonstrated that individuals with SCA showed reduced practice benefits on both sequence and random blocks (Tzvi et al., 2017), similar to the deficits we observed for both repeating and non-repeating sequences, respectively.

Note that the literature on motor sequence learning in PD is not unequivocal: While the majority of studies indicate impaired sequencing skill in PD, a group of studies still opposes this conclusion (for a review see (Ruitenberg et al., 2015)). Previous studies have produced mixed results which may suggest that the involvement of the BG is selective and depends on the specific learning conditions (Ruitenberg et al., 2015, 2016; Shin & Ivry, 2003b). For example, in studies utilizing a modified version of the SRTT, PD patients were less efficient in learning random non-repeating sequences, but had no impairment in learning repeated sequences (Muslimović et al., 2007; Ruitenberg et al., 2016; Tremblay et al., 2010). Thus, in line with our results, while we observed deficits in the algorithm-based learning, memory-based learning may be spared in PD.

## Discussion

In this paper we discribe a novel protocol for online neurop sychological testing. PONT was designed to take advantage of features that have motivated many experimental psychologists to move to on-line studies over the past decade, yet it is tailored to address the unique demands of patient-based research. The main focus of this paper was to outline the key steps required for this approach, with our report of the results from a sequence learning included to provide a concrete example of the application of PONT.

PONT was developed to address five main challenges facing researchers who conduct neuropsychological studies. Perhaps the most profound limiting factor that this valuable approach addresses is access to a targeted population (see Figure 4), be it a disorder with a relatively high prevalence rate in the population (e.g., 0.3% for PD) or one that is rare (e.g., 0.04% for SCA). Not only can it be difficult to recruit these individuals, but the patients themselves may have limited time and energy given their neurological condition. As a result, it can take a long time to complete a single experiment (e.g., 1-2 years), a problem that is magnified for a project requiring multiple experiments. Indeed, neuropsychological papers tend to involve just a single patient group and single experiment. PONT provides a protocol that makes data collection extremely efficient. In the first 10 months following the launch of PONT, we were able to complete six experiments, with each having around 20 participants per group. Moreover, as our patient participant pool grows, we should be in a position to run studies with much larger sample sizes, something that is relatively rare in the cognitive neuroscience literature but desirable for looking at factors underlying heterogeneity in performance within a particular group (e.g., due to genetic subtype, pattern of pathology).

**Figure 4.**
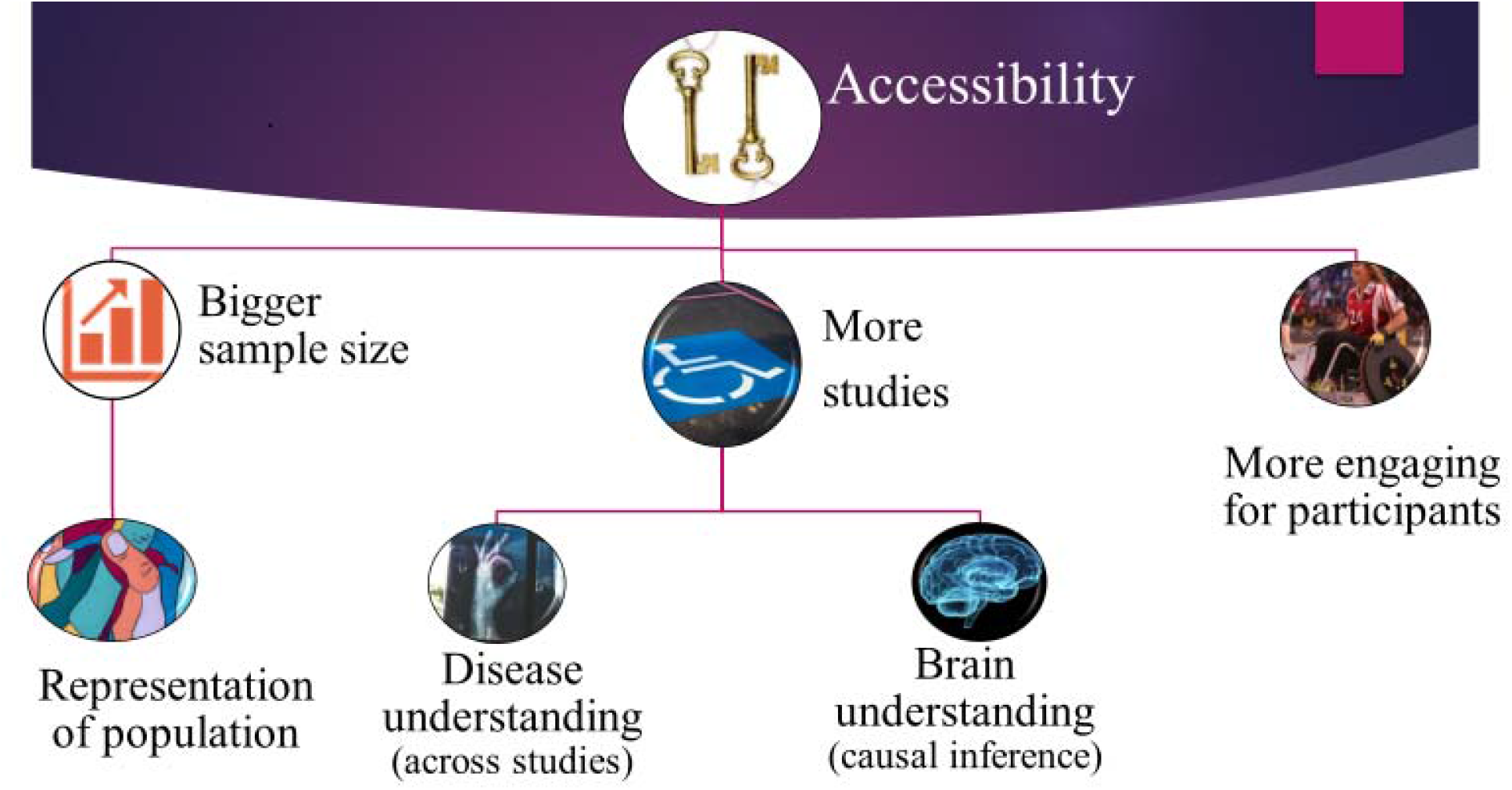
Advantages of the online protocol for neuropsychological research.

Second, recruitment for laboratory-based experiments usually comes from a local community. As such, the sample may be small and not representative of the population (see Figure 4). For example, our PD sample from the Berkeley community tend to be highly educated compared to the general PD population in the USA. In addition, if the study requires a relatively homogenous pathology or genetic subtype (e.g., a particular variant of SCA), the sample size likely to be small (e.g., 8 participants). By casting the recruitment net across the entire country (or, as we envision, internationally), PONT will be ideal for running experiments with larger and more diverse samples that will better represent the general population.

Third, while we developed PONT to facilitate our research program on subcortical contributions to cognition, this protocol can be readily adopted for different populations. It can be readily adopted for disorders such as Alzheimer’s disease, stroke, and dyslexia which are associated with support groups or online social networks. We anticipate recruitment will be more challenging for any individual lab when these points of contact are absent. However, PONT can provide a common test protocol to support multi-lab collaborative operations, an alternative way to increase the participant pool that may be especially useful for studies aimed at more targeted populations (e.g., medulloblastoma, non-fluent aphasia). As such, PONT provides a valuable approach to conduct neuropsychological research to explore a wide range of brain regions and neurological disorders.

An important benefit of PONT is that it is user-friendly for the participants. The participants complete the experiment at home, choosing a time that fits into their personal schedule. As we have learned from our experience over the past three decades working with SCA and PD participants, arranging transportation to and from the lab can present a major obstacle, especially when the travel is to participate in a study that offers no direct clinical benefit to the participants. The participants also seem to enjoy the challenge of their “assignment,” although they, like our college-age participants may complain about the repetitive nature of a task when hundreds of trials are required to obtain a robust data set. Table 2 provides a sample of the comments we have received in our post-session surveys. Although we had embarked on the PONT project prior to the onset of the COVID-19 pandemic, the timing was fortuitous, allowing us to continue and ramp up our neuropsychological testing, allowing vulnerable populations to participate in studies in a safe manner. PONT can serve as an “immune system” for neuropsychological research development––protecting research development even when patients’ accessibility is more limited than ever.

**Table 2.**
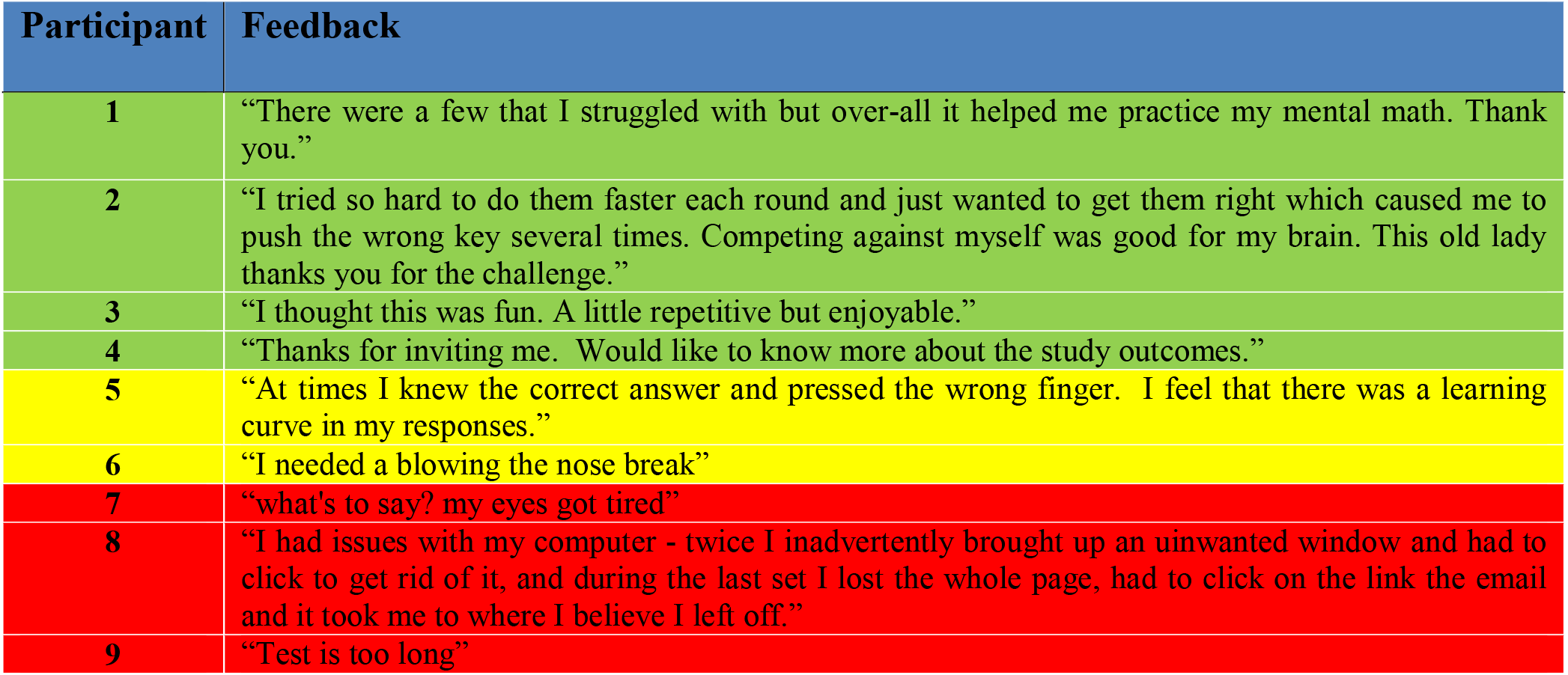
A sample of the Participants’ feedback after completing the online experiments. Green – positive feedback, yellow – neutral, red – negative feedback.

| Participant | Feedback |
| --- | --- |
| 1 | "There were a few that I struggled with but over-all it helped me practice my mental math. Thank you." |
| 2 | "I tried so hard to do them faster each round and just wanted to get them right which caused me to push the wrong key several times. Competing against myself was good for my brain. This old lady thanks you for the challenge." |
| 3 | "I thought this was fun. A little repetitive but enjoyable." |
| 4 | "Thanks for inviting me. Would like to know more about the study outcomes." |
| 5 | "At times I knew the correct answer and pressed the wrong finger. I feel that there was a learning curve in my responses." |
| 6 | "I needed a blowing the nose break" |
| 7 | "what's to say? my eyes got tired" |
| 8 | "I had issues with my computer - twice I inadvertently brought up an unwanted window and had to click to get rid of it, and during the last set I lost the whole page, had to click on the link the email and it took me to where I believe I left off." |
| 9 | "Test is too long" |

Lastly, PONT provides considerable benefit in terms of reduced economic and environmental cost. Setting aside the hard-to-calculate benefits from reduced travel and reductions in personnel time, the savings are substantial when just considering participant costs. For our in-lab studies, we typically conduct two 35-min experiments, sandwiched around a 30 min break. When we factor in compensation for travel time, the cost tends to come to around $100/session. The same amount of data can be obtained for approximately $40 over two PONT sessions, with the added benefit that potential fatigue effects are reduced given that each session is run on a different day.

Although there are many advantages to this online approach, there are also some notable limitations that need to be taken into account. First, the procedure for online experiments is likely to be less standardized across participants. A major source of variability comes about because participant uses his/her own computer and response device. These are, of course, standardized for in-lab studies, including positioning the participants such that the visual displays are near-identical. It may be possible to impose some degree of control on viewing angle by specifying how the participants’ position themselves, but this is likely going to be challenging when the participants are self-administering the experimental task.

Second, online experiments are unlikely to be well-suited for all domains of study. For example, given the variability in hardware, it would be difficult to run experiments that require precise timing; for example, studies in which stimuli are presented for a short duration followed by a mask. Moreover, we have avoided studies that use auditory stimuli since computer speakers are so variable and can be of poor quality, and it is not possible use tactile or olfactory stimuli.

Third, the use of an automated, self-administered system comes with a cost. In our in-person studies, it is not uncommon for a participant to misunderstand the instructions or need extra practice when they find a particular task difficult. These situations can be readily identified and addressed when an experimenter is present. Moreover, a participant can get discouraged if they find the task too challenging or have an error rate higher than their personal “standard.” An encouraging experimenter can help ensure motivation remains high. We incorporate various feedback messages to keep the participants’ morale and motivation high, but an online system will be less flexible than in-person testing (which could be seen as a positive for maintaining test uniformity). We imagine there will be some experiments that may require the virtual presence of an experimenter. We have also conducted post-experiment live check-ins with some participants to both maintain a human touch and get feedback on how we can make the experience more enjoyable and beneficial. Inspired by the feedback, we have started providing periodic newsletters to the participants, describing recent findings from both basic and translational studies (https://mailchi.mp/ab940c63fa5c/newsletter-by-the-cognac-lab-at-uc-berkeley-the-neuroscience-of-ataxia-3556742?e=ae8c913a37).

Lastly, but of critical importance, is concern about the quality of the data when testing is self-administered and the environment may be conducive to distraction (e.g., from a TV in the background or attention-grabbing text alerts). Although the Covid-19 pandemic precluded a direct comparison of in-person and online testing with the PONT protocol, the results from the sequence learning study seem similar to those published from in-person experiments. Several studies have made more systematic comparisons of online and in-person experiments (Barnhoorn, Haasnoot, Bocanegra, & van Steenbergen, 2014; Simcox & Fiez, 2014). Overall, the results are encouraging in that, while the data may be noisier, the general patterns are similar. We note that this is especially true with large sample sizes, something that is unlikely to be true in many neuropsychological studies. One recent review provides recommendations for improving data quality that are certainly appropriate for PONT (Grootswagers, 2020): Keep the experiments as short as possible, provide reasonable compensation, and make the tasks as engaging as possible.

## Conclusion

The emergence of online protocol s has provided behavioral scientists with the opportunity to conduct studies that involve large and diverse samples, while being cost efficient. The PONT protocol described in this paper describes how this general approach can be adopted to meet the challenges associated with neuropsychological testing. Currently, our work is limited to individuals with ataxia and PD, allowing us to expand our research program on subcortical contributions to cognition. The materials provided with this paper can be readily adopted by researchers working with any patient population, especially when recruitment can be conducted via support groups, web-based groups, or through collaborations across multiple labs/clinics. In terms of the latter, we see PONT as a fertile tool to support multi-national collaborative research operations. We expect PONT will significantly increase the sample size, the number of studies conducted, and the overall pace of neuropsychological research. As such, it offers a powerful tool for this field, one that has and will continue to yield fundamental insights into brain-behavior relationships.

## Data Availability

Materials are available at https://osf.io/fktn9/

## Data Availability

Materials are available at https://osf.io/fktn9/

## Acknowledgments

We thank Sharon Binoy, Nandita Radhakrishnan, Rachel Woody, Sidra Seddiqee, and Sravya Borra for their assistance. This research was supported by funding from the National Institute of Health (NS116883).

## Open Practices Statement

Materials and data are available at https://osf.io/fktn9/.

## Appendix

Full weblink to materials for running the PONT protocol is accessible in open science framework - https://osf.io/fktn9/.

